# The Association between Sleep Quality and Type Two Diabetes at 20 Year Follow-up in the Southall And Brent REvisited (SABRE) Cohort: a Tri-ethnic Analysis

**DOI:** 10.1101/2020.08.23.20157073

**Authors:** Zhen L. Ong, Nishi Chaturvedi, Therese Tillin, Caroline Dale, Victoria Garfield

## Abstract

**Objective:** The risk of developing type 2 diabetes associated with poor sleep quality is comparable to that of traditional risk factors (e.g. overweight, physical inactivity). In the United Kingdom, these traditional risk factors could not explain the two to three-fold excess risks in South Asian and African Caribbean men compared to Europeans. This study investigates the (i)the association between mid-life sleep quality and later-life type 2 diabetes risk and (ii)a potential modifying effect of ethnicity.

**Research Design and Methods:** The Southall and Brent REvisited (SABRE) cohort comprised Europeans, South Asians, and African Caribbeans (median follow-up = 19 years). Complete case analysis was performed on 2190 participants without diabetes at baseline (age= 51.7± 7SD). Competing risks regressions were used to estimate the hazard ratios (HRs) of developing type 2 diabetes associated with four self-reported baseline sleep exposures (difficulty falling asleep, early morning waking, waking up tired and snoring) while adjusting for confounders. Modifying effects of ethnicity were analysed by (i) testing for interactions and (ii) performing ethnicity-stratified analysis.

**Results:** Snoring was strongly associated with increased type 2 diabetes risk but only among South Asians in a fully-adjusted model (HR 1.42, 95%CI=1.08-1.85, P=0.011). Our results revealed no elevated risk for any of the sleep exposures across all three ethnic groups.

**Conclusions:** The association between snoring and type 2 diabetes appeared to be modified by ethnicity, with South Asians at greatest risk.

---

There is growing evidence for an association between sleep duration or quality and type 2 diabetes. Compared to sleep duration, fewer studies have explored the potential long-term effect of the equally important aspect of sleep quality on type 2 diabetes (1). While sleep duration refers to the number of hours asleep, sleep quality refers to difficulty initiating sleep, difficulty maintaining sleep (e.g. sleep-disordered breathing or uncontrollable limb movements), and the subjective feeling of being rested (2-4). Obstructive sleep apnoea (OSA) is a severe but relatively common form of sleep-disordered breathing (5), characterised by loud snoring, breathing cessation and repeated nocturnal awakenings. A recent meta-analysis of prospective cohort studies revealed that the adjusted pooled relative risks of developing type 2 diabetes associated with difficulty initiating sleep, difficulty maintaining sleep and OSA were 1.55, 1.72 and 1.49 respectively. These effect sizes were comparable to well-established type 2 diabetes risk factors, and only marginally smaller than having a family history of diabetes or overweight, but greater than being physically inactive (6).

Even fewer studies have included ethnic minorities (5,7), specifically South Asians and African Caribbeans, despite their two-to three-fold increased risk of type 2 diabetes compared to those of European origin (8). These excess risks appear only partially due to traditional biological and environmental risk factors(8). Given the potential social patterning of sleep (9) and that OSA may be more prevalent among ethnic minorities (10), we hypothesised that there would be (i) a deleterious association between specific sleep quality measures (difficulty falling asleep, early morning waking, waking up feeling tired, and snoring) and type 2 diabetes risk, and (ii) that these associations would be stronger in people of South Asian or African Caribbean origin. Since sleep quality is potentially modifiable, understanding its role in the development of type 2 diabetes may offer new opportunities for the prevention of type 2 diabetes.

## Research Design and Methods

### Study Design and Population

Southall And Brent REvisited (SABRE) (2008-12) is a multi-ethnic community-based prospective cohort composed of older Europeans, South Asians, and African Caribbeans from London. Median follow-up period was 19 (IQR=15, 20) years. The cohort was specifically set up to examine ethnic differences in cardiometabolic disorders. At baseline (1988-1991), there were more Europeans (48.8%) than South Asians (38.8%) and African Caribbeans (12.4%) (11). Participants were aged between 40-69 years old and 75% were men (by initial study design) (8). Participants’ ethnicity was initially determined by interviewers based on grand-parental origin and confirmed by participants (8). All South Asians and African Caribbeans were first-generation migrants. The majority of African Caribbeans originated from the Caribbean (91.5%) and the rest from West Africa (8). South Asian participants consisted of Punjabi Sikhs (52%), Gujarati or Punjabi Hindus (20%), Muslims (15%) and other South Asians (15%). Survivors at follow-up were between 57-90 years old. A detailed cohort profile (11) and information on follow-up (8) have been published elsewhere. The present study used data from baseline and 20-year follow-up.

Participants gave informed consent. The baseline study was approved by Ealing, Hounslow and Spelthorne, Parkside, and University College London research ethics committees. The follow-up study was approved by St. Mary’s Hospital Local Research Ethics Committee (reference 07/HO712/109).

All Indian Asians and African Caribbeans were first-generation migrants. Ethnicity was interviewer-recorded based on parental origin and appearance and was subsequently confirmed by participants. Indian Asians originated from the Indian subcontinent. African Caribbeans originated from the Caribbean (91.5%) or from West Africa.

### Measures

#### Exposure: Sleep Quality

At baseline, participants answered four questions on sleep quality, including whether they (i) had difficulty falling asleep, (ii) woke up too early, (iii) felt tired upon waking up and (iv) snored in the past 30 days. The first three questions were adapted from the Jenkin’s Sleep Questionnaire (JSQ) (12), which is a brief, reliable and widely used sleep disturbance questionnaire validated among air traffic controllers and patients recovering from cardiac surgery. All responses were recoded as binary variables (yes=1 or no=0). The snoring variable was dichotomised at ‘often’ and ‘almost always’ as ‘yes’, whereas ‘occasionally’ and ‘often’ were coded as ‘no’. Snoring was used as a proxy marker for OSA, which is a more severe form of sleep-disordered breathing.

#### Outcome: Incident Type 2 Diabetes

Because sleep disturbances may be a symptom of type 2 diabetes, participants with diabetes at baseline were excluded to ensure that sleep quality exposures preceded any type 2 diabetes outcome. *Baseline* type 2 diabetes was ascertained by (i) self-report of doctor diagnosis or (ii) receipt of anti-diabetes medications (iii) fasting blood glucose ≥7.0 mmol/L or post load glucose after an oral glucose tolerance test (OGTT) ≥ 11mmol/L according to the World Health Organisation (13) criteria.

For *incident* type 2 diabetes status, participants were directly followed-up using (i) primary-care medical record review of diagnosis or prescribed antidiabetic medication, (ii) self-completion questionnaires of physician diagnosis in addition to the year of diagnosis or named antidiabetic medication, and (iii) OGTT ≥ 11mmol/L at follow-up (11). Incident diabetes status of survivors obtained by direct follow-up was readily available as binary outcomes in the dataset owing to previous work done by Tillin et al., 2013.

Participants who had died due to type 2 diabetes between baseline and follow-up (before 2011) but did not have a recorded year of diagnosis were excluded from the study sample, as this information was necessary for competing risks regression modelling.

#### Covariates

A range of baseline-measured covariates were adjusted for, grouped by (i) demographic factors (age, sex, ethnicity and socioeconomic position (SEP) as measured by years of education), (ii) self-reported health behaviours (smoking status and total weekly leisure physical activity), and (iii) general adiposity (BMI). These covariates were established a priori risk factors for type 2 diabetes (8,14). Our study controlled for general adiposity instead of central adiposity (using waist-to-hip ratio) as several studies suggest that general adiposity may be more important in impacting sleep quality, particularly for OSA (15–17).

### Statistical Analyses

Statistical analyses were carried out using RStudio version 1.1.456 and STATA.

This study used a complete case analysis (CCA) to ensure comparability across different models. This study excluded participants with diabetes at baseline, those lost to follow-up for diabetes status (LTFU), and participants with missing data on any covariates specified in the models. The descriptive analyses were performed on n=2194 participants. However, the competing risks regression analysis was conducted on n= 2190 participants. The authors were unable to harmonise these figures as COVID-19-related government regulations prevented access to restricted data held at the workplace.

### Descriptive Statistics

Baseline characteristics of participants were presented for all covariates, grouped by incident type 2 diabetes status. Results are expressed as geometric mean±SD for continuous variables and frequency with percentages for categorical variables. In this study, all formal tests of differences between groups were conducted using one-way analysis of variance (ANOVA) for continuous variables and Pearson’s chi-squared (χ^2^) tests for categorical variables. T-tests were not used in this study.

The patterns of sleep quality exposures were also examined by ethnicity and incident type 2 diabetes status.

### Competing Risks Regression

Competing risks hazard regression models were used to calculate (sub) hazard ratios (HRs) and 95% confidence intervals (CI) of developing type 2 diabetes for the four sleep quality exposures specified within the same model. For each sleep quality exposure, those who reported no sleep problems formed the reference group.

The outcome for the survival model was time from baseline until the development of type 2 diabetes or censoring. Death from other causes than type 2 diabetes was considered as a competing event. Participants without death notification were censored at the end of the follow-up time (2011).

Three hierarchical competing risk models were specified. Model 1 controlled for demographic factors (age, sex, ethnicity and education); Model 2 additionally adjusted for health behaviours (smoking status and physical activity); Model 3 additionally adjusted for general adiposity (BMI).

### Modifying effects of Ethnicity

The potential modifying effects of ethnicity on the association between poor sleep quality and incident type 2 diabetes were initially explored by adding an **interaction term** for *ethnicity*sleep quality exposures* in the fully adjusted model (Model 3). However, these interaction effects were tested for each sleep variable one at a time. Any interaction terms below the alpha threshold of 0.10 were taken forward by running the models within **ethnicity-stratified samples**, in which the conventional alpha of 0.05 was then applied.

### Attrition

More than one-third of participants without diabetes at baseline were lost to follow-up (LTFU) on incident type 2 diabetes status despite extensive efforts for tracing and follow-up. Missingness on all other variables of interest was negligible (<5%). Baseline characteristics of those LTFU were compared to the CCA sample.

## Results

### Baseline Characteristics

**Table 1** shows the baseline characteristic of participants with complete data for all the covariates, stratified by incident diabetes status. Among the 2194 complete cases, 488 people (22%) had developed type 2 diabetes at 20-year follow up. Only 14% of the Europeans developed type 2 diabetes, in contrast with 33% of South Asians and 30% of African Caribbeans. Type 2 diabetes incidence was also associated with nearly all model covariates, except for socioeconomic position.

**Table 1.** —Baseline characteristics of participants by incident type 2 diabetes status, complete cases (n=2194)

| Baseline characteristics | Incident T2D<br>(N=488) | No T2D<br>(N=1706) | P-value<br>for trends |
| --- | --- | --- | --- |
| **Age (years)** | 50.8 (6488).4) | 52.2 (7.1) | **<0.001\*\*\*** |
| **Sex (proportion male)** | 397 (81) | 1309 (77) | **0.035\*** |
| **Ethnicity** |  |  | **<0.001\*\*\*** |
| European | 170 (35) | 1031 (60) |  |
| South Asian | 233 (48) | 473 (28) |  |
| African Caribbean | 85 (17) | 202 (12) |  |
| **SEP (Years of education)** | 11.4 (3.2) | 11.2 (3.1) | 0.291 |
| **Physical Activity (MJ/wk)** | 10.2 (6.8) | 11.1 (7.2) | **0.017\*** |
| **Smoking Status** |  |  | **0.002\*\*** |
| Never smoker | 276 (57) | 838 (49) |  |
| Ex-smoker | 94 (19) | 453 (27) |  |
| Current smoker | 118 (24) | 415 (24) |  |
| **BMI (kg/m<sup>2</sup>)** | 27.62 (4.2) | 25.62 (3.7) | **<0.001\*\*\*** |

BMI, body mass index; SEP, socioeconomic position; T2D, type 2 diabetes;
P: \*\*\*<0.001, \*\*<0.01, \*<0.05

### Pattern of Sleep Quality Exposures

Overall, early morning waking was the most commonly reported sleep problem (40%), followed by tiredness upon waking (38%), snoring (36%) and difficulty falling asleep (19%). For all ethnicities combined, snoring prevalence was found to be different between type 2 diabetes cases and non-cases (43% and 36% respectively). This difference seemed to be driven by snoring patterns among South Asians **(Figure 1)**. Snoring was reported by 47% of South Asians who developed type 2 diabetes, in contrast with 33% by non-cases. Inter-ethnic differences were also found for difficulty sleeping and early morning waking; African Caribbeans tended to have greater difficulty falling asleep; South Asians tended to wake up too early.

**Figure 1.**
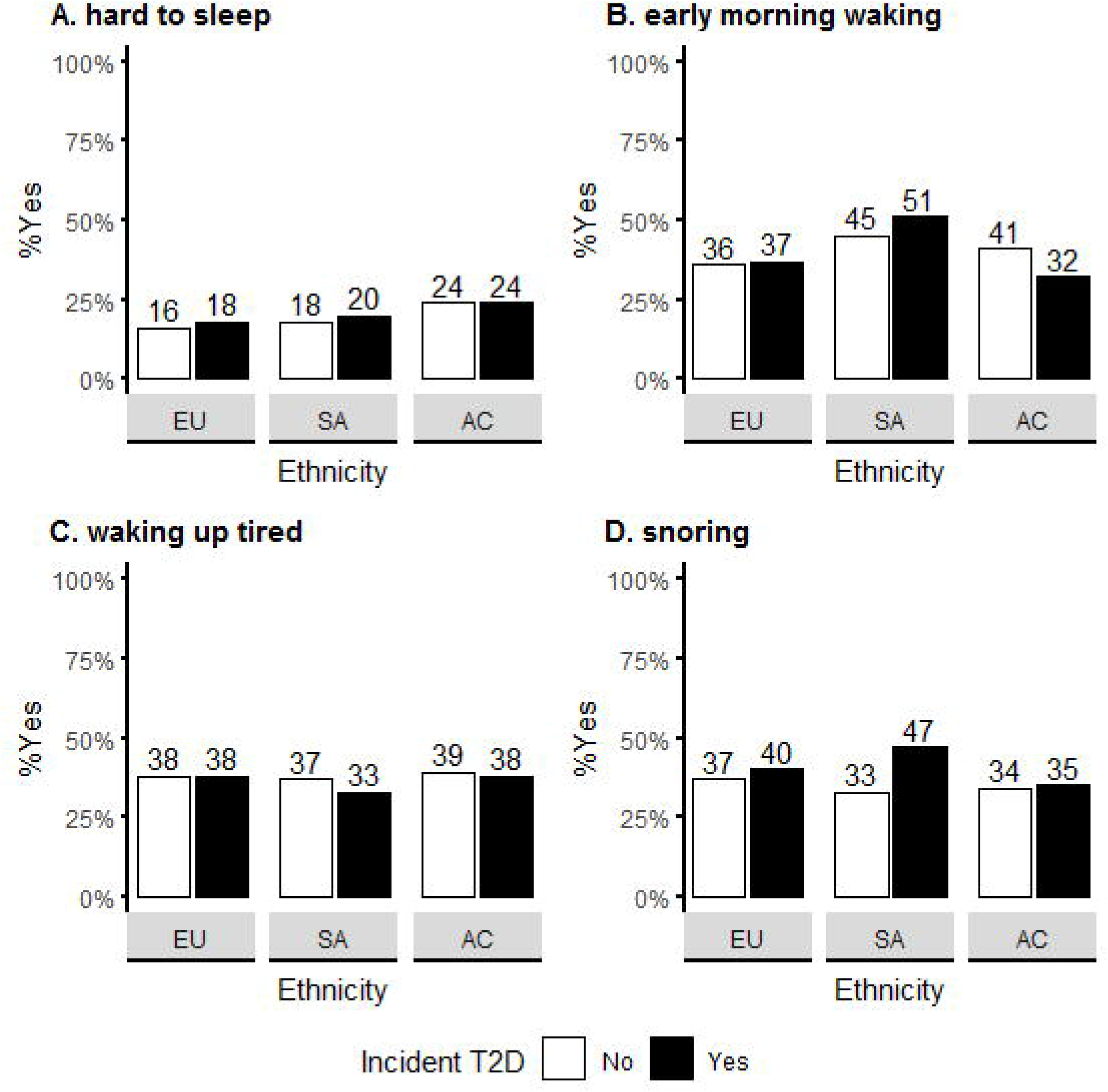
— Prevalence of sleep problems by ethnicity and incident type 2 diabetes status. T2D, type 2 diabetes; EU, Europeans; SA, South Asians; AC, African Caribbeans. Y-axis represents the percentage of “yes” respondents for the sleep quality exposures. Sample sizes (% of complete case sample): A. n=406(19%); B. n=887(40%); C. n=836(38%); D. n=1370(62%)

### Modifying Effects of Ethnicity

Applying an alpha of 0.10 to the **interaction** effect models, results suggested a modifying effect of ethnicity on the association between snoring and incident type 2 diabetes risk only (P=0.056). For snoring, results show a modifying effect of being of South Asian origin compared to Europeans in the fully adjusted model (P=0.048).

**Figure 2** shows the HRs and 95% CIs of developing type 2 diabetes at 20-year follow-up for snoring, **stratified by ethnicity** in complete cases (n=2190). A distinctly stronger link between snoring and incident type 2 diabetes was found for South Asians, as compared to Europeans. South Asians who snored were at greater risk of developing type 2 diabetes (HR 1.42, 95%CI=1.08-1.85, P=0.011), compared to their non-snoring counterparts which was not due to differences in measured covariates (Model 3). This association was not apparent among Europeans (HR 0.91, 95%CI =0.67-1.23, P=0.534) or African Caribbeans (HR 0.93, 95%CI =0.59-1.46, P=0.742) in minimally- or fully-adjusted models.

**Figure 2.**
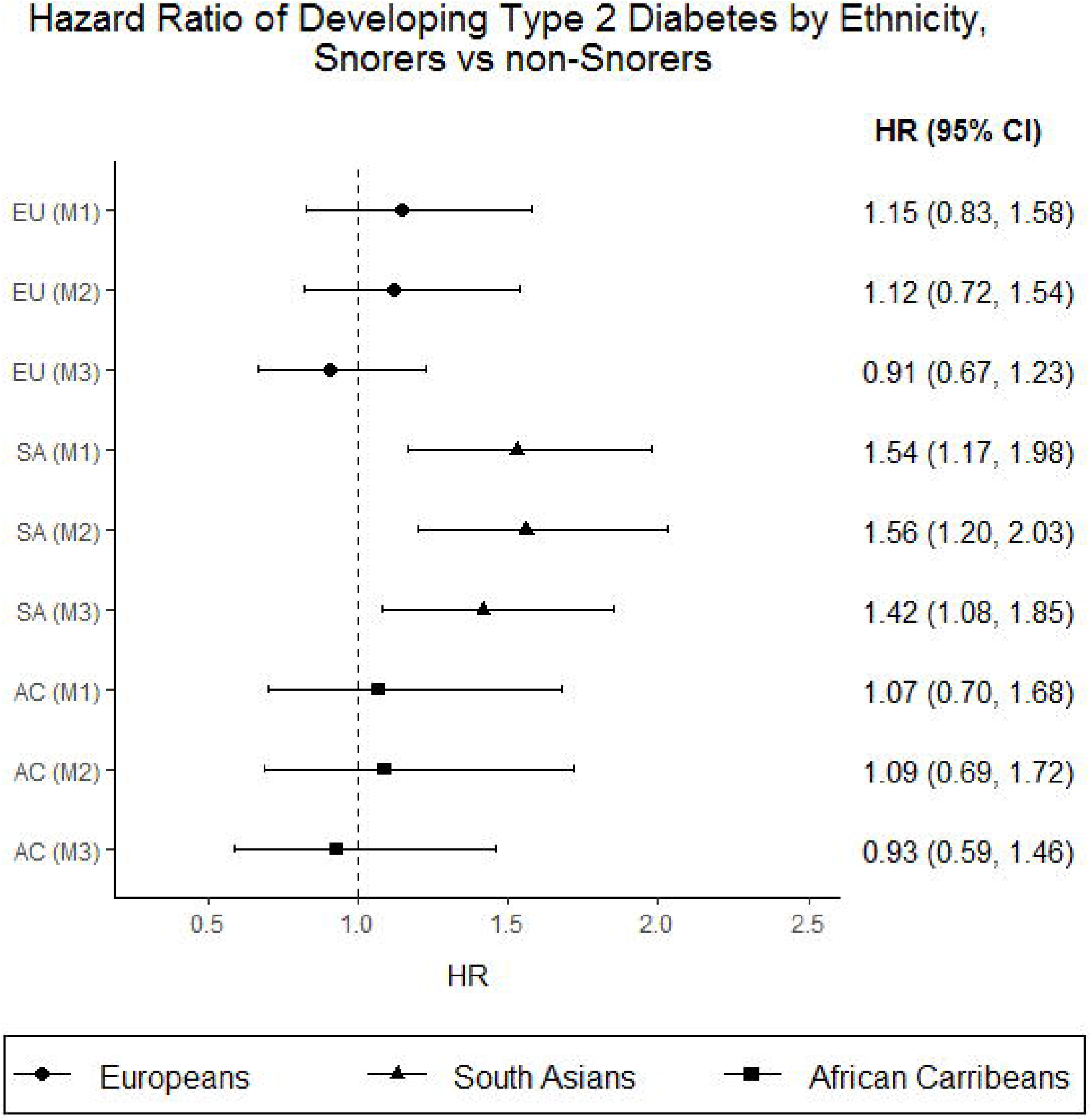
— Ethnicity-stratified analysis for snoring, complete-case analysis (n=2190). Circles = Europeans; triangles = South Asians; squares = African Carribeans BMI, Body Mass Index; EU, Europeans; HR, hazard ratio; SA, South Asians; AC, African Caribbeans; M1-M3 =Models 1-3. Model 1: basic model, adjusted for age, sex and socioeconomic position Model 2: model 1 plus physical activity and smoking status Model 3: model 2 plus BMI

**Table 2** shows the results of the ethnicity-stratified analysis for difficulty falling asleep, early morning waking and feeling tired upon waking up. For difficulty falling asleep and early morning waking, there were no apparent association with the risk of developing type 2 diabetes in all models. Contrary to our hypotheses, there was a relatively strong negative association between waking up tired and incidence of type 2 diabetes (0.72, 0.54-0.97, P=0.029) (Model 3) among South Asians.

**Table 2.**
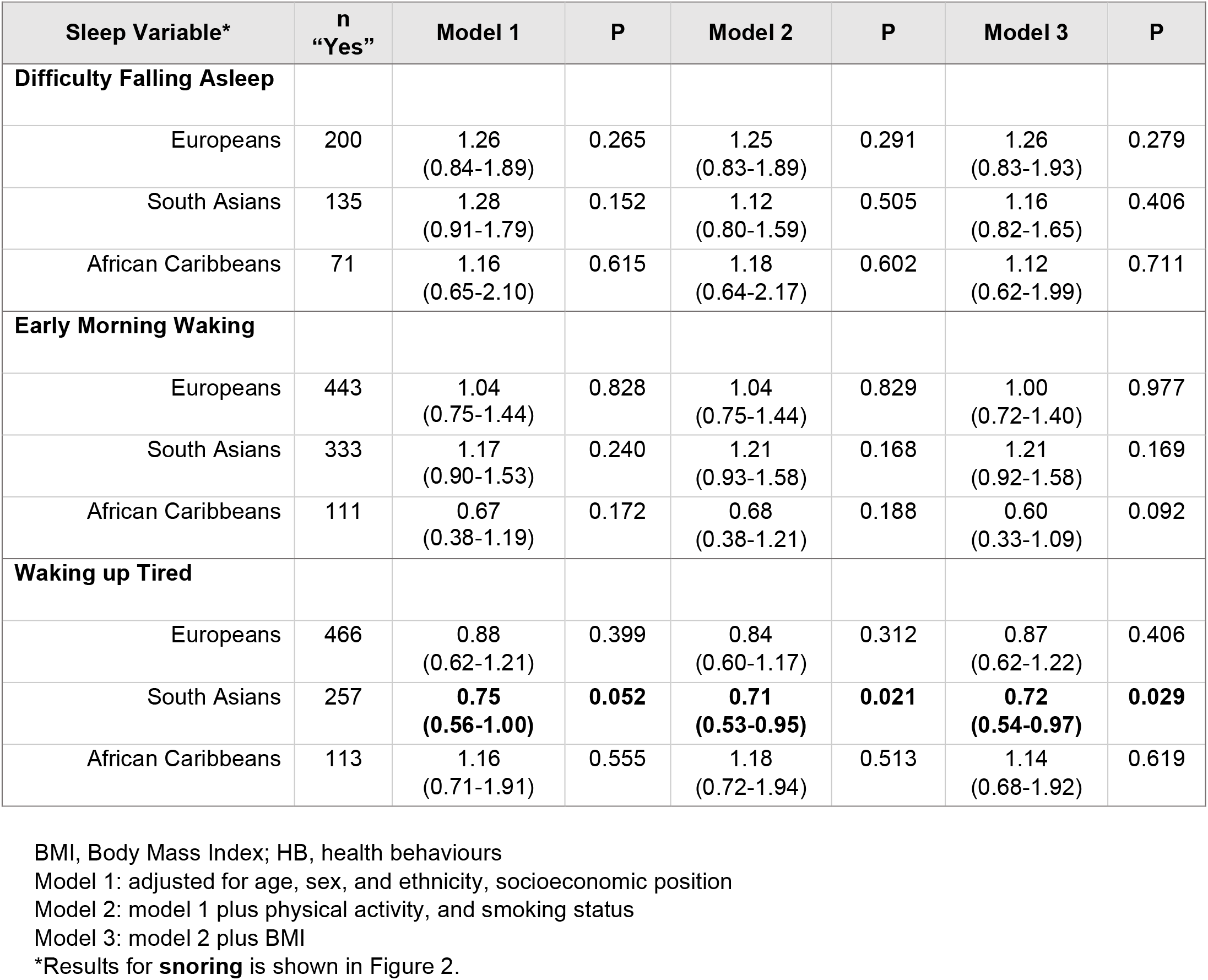
—Ethnicity-stratified analysis of associations between sleep quality exposures and incident type 2 diabetes reported in hazard ratios (95%CI), complete case analysis (n=2190)

| Sleep Variable* | n<br>“Yes” | Model 1 | P | Model 2 | P | Model 3 | P |
| --- | --- | --- | --- | --- | --- | --- | --- |
| **Difficulty Falling Asleep** |  |  |  |  |  |  |  |
| Europeans | 200 | 1.26<br>(0.84-1.89) | 0.265 | 1.25<br>(0.83-1.89) | 0.291 | 1.26<br>(0.83-1.93) | 0.279 |
| South Asians | 135 | 1.28<br>(0.91-1.79) | 0.152 | 1.12<br>(0.80-1.59) | 0.505 | 1.16<br>(0.82-1.65) | 0.406 |
| African Caribbeans | 71 | 1.16<br>(0.65-2.10) | 0.615 | 1.18<br>(0.64-2.17) | 0.602 | 1.12<br>(0.62-1.99) | 0.711 |
| **Early Morning Waking** |  |  |  |  |  |  |  |
| Europeans | 443 | 1.04<br>(0.75-1.44) | 0.828 | 1.04<br>(0.75-1.44) | 0.829 | 1.00<br>(0.72-1.40) | 0.977 |
| South Asians | 333 | 1.17<br>(0.90-1.53) | 0.240 | 1.21<br>(0.93-1.58) | 0.168 | 1.21<br>(0.92-1.58) | 0.169 |
| African Caribbeans | 111 | 0.67<br>(0.38-1.19) | 0.172 | 0.68<br>(0.38-1.21) | 0.188 | 0.60<br>(0.33-1.09) | 0.092 |
| **Waking up Tired** |  |  |  |  |  |  |  |
| Europeans | 466 | 0.88<br>(0.62-1.21) | 0.399 | 0.84<br>(0.60-1.17) | 0.312 | 0.87<br>(0.62-1.22) | 0.406 |
| South Asians | 257 | **0.75**<br>**(0.56-1.00)** | **0.052** | **0.71**<br>**(0.53-0.95)** | **0.021** | **0.72**<br>**(0.54-0.97)** | **0.029** |
| African Caribbeans | 113 | 1.16<br>(0.71-1.91) | 0.555 | 1.18<br>(0.72-1.94) | 0.513 | 1.14<br>(0.68-1.92) | 0.619 |

BMI, Body Mass Index; HB, health behaviours
Model 1: adjusted for age, sex, and ethnicity, socioeconomic position
Model 2: model 1 plus physical activity, and smoking status
Model 3: model 2 plus BMI
\*Results for **snoring** is shown in Figure 2.

### Attrition

Participants who were lost to follow-up (LTFU) tended to be older, have fewer years of education, less physically active and have higher BMI. Males were less likely to be lost to follow-up (CCA=78% vs. LTFU=72%).

## Conclusions

Results show that snoring was strongly associated with incident type 2 diabetes, observed only in South Asians but not in Europeans and African Caribbeans. Difficulty falling asleep, early morning waking, and feeling tired upon waking up were not associated with elevated type 2 diabetes risk across all ethnic groups.

Our null findings for difficulty falling asleep and difficulty maintaining sleep (measured here as “early morning waking”) were not comparable to results from a recent meta-analysis (6). Some study-specific characteristics may explain these results. Studies included in the meta-analysis typically involved over 2000 ethnically homogenous groups from wealthy countries. Although most studies included in recent meta-analyses employed similar self-reported sleep quality questions, biased interpretation due to cultural differences may exist. For instance, African Americans were less likely to complain of a “sleep problem” but more likely to report taking over 30 minutes to fall asleep, which is the typical clinical cut-off (18,19). If a similar bias exists in other ethnic groups, then insomnia-symptoms may be underestimated, potentially explaining the lack of adverse effects in this study. Moreover, the Jenkin’s Sleep scale has not been cross-culturally validated. The questions may be more appropriate for Western monophasic sleep cultures (one sleep session at night) but less suited for the siesta sleep culture (daytime napping and nocturnal sleep) common in Caribbean communities (20). Future sleep research should use cross-culturally validated sleep questionnaires or multigroup analysis to ensure measurement invariance across ethnic groups.

The strong association found for snoring among South Asians is comparable to the pooled unadjusted relative risk for OSA in the same meta-analysis (6). The two-fold increased type 2 diabetes risk associated with snoring observed only in South Asians represents a novel finding. In an attempt to extend previous work on ethnic differentials in type 2 diabetes risk (8), our findings suggest that snoring could play a more prominent role in type 2 diabetes pathogenesis in South Asians. While this study did not adjust for an identical set of covariates as the study by Tillin and colleagues (8), uncovering the causes of the South Asians’ heightened type 2 diabetes vulnerability associated with snoring is an important research implication. Our study could not measure objectively diagnosed OSA and we did not capture the intensity of snoring, which might be informative in suggesting presence of OSA (21) Nevertheless, previous studies have found that the progression of increased apnoea/hypopnoea index (an indicator of the presence and severity of OSA) among primary snorers and untreated patients with mild to moderate OSA were significantly associated with length of time and increased weight (22). Furthermore, the prevalence and severity of OSA among South Asians are higher than Europeans for the same level of BMI (23), which could be partially attributed to a greater propensity among South Asians to visceral adiposity at a lower BMI compared to white Europeans (24).

Short-term laboratory studies have found several mechanisms linking poor sleep quality in general to impaired glucose metabolism, upregulation of appetite and similar-levels or reduced total energy expenditure due to daytime fatigue, predisposing individuals to weight gain and risk of type 2 diabetes (25,26). Of note, three nights of laboratory-controlled sleep fragmentation (reducing slow-wave sleep, the deepest sleep stage) without changes in total sleep duration, showed a 25% decreased insulin sensitivity without a compensatory increase in insulin secretion among healthy volunteers (27). This effect was comparable to a difference in type 2 diabetes risk associated with being eight to 13 kilograms heavier (28). This suggests an adverse effect of poor sleep quality on the pathogenesis of type 2 diabetes independently of total sleep duration. For OSA specifically, mechanistic explanations mainly involve (i) sleep fragmentation/ reduced slow wave sleep due to repeated night time awakening, (ii) intermittent hypoxia (periodic deoxygenation and reoxygenation of blood due to recurrent airway blockage), and (iii) reduced total sleep duration. These states may predispose individuals to glucose intolerance and insulin resistance through multiple pathways, including systemic inflammation and increased oxidative stress, ultimately increasing type 2 diabetes risk (25). Moreover, the association between snoring and type 2 diabetes risk independent of adiposity was consistent with findings from a meta-analysis of prospective cohorts which examined OSA (29). However, adjusting for adiposity contributed to the greatest attenuation in type 2 diabetes odds in this study, which suggested a partial mediation or confounding effect. Obesity is a prominent shared risk factor for OSA and type 2 diabetes. Fat deposition surrounding the neck, chest, and abdomen (30) may compromise airway space and collapsibility, predisposing individuals to OSA. Yet, frequent snoring has also been found to be independently associated with glucose intolerance in lean adults (31). Without information regarding the temporality of adiposity and OSA onset, it was unclear from this study whether adiposity acted as a confounder or a mediator. Future research to disentangle the pathogenesis of type 2 diabetes due to obesity and/or OSA may benefit from a life-course perspective.

We also acknowledge that our study had several limitations. We only used sleep quality data at one time point in 20-years and as such could not capture the cumulative exposures of sleep problems and changes over time. Ageing is associated with decreased sleep quantity and quality (32,33), and more daytime napping (34). Retirement may have changed participants’ sleep quality if sleep disturbances such as insomnia were work-related (e.g. shift-work, work-related stressors). More free time post-retirement may also allow for longer sleep or napping to compensate for poor sleep quality. Indeed, laboratory experiments have found that recovery sleep following a time of restricted sleep could partially reverse disruption in glucose metabolism over the short term (35) but long-term impacts of improved sleep require further investigation. For migrant groups, poor sleep quality caused by acculturative stress might also improve over time upon successful integration to the host society (36). Another limitation common to observational studies includes residual confounding, particularly for family history of diabetes, dietary patterns (e.g. hypercaloric diets, caffeine intake) (37) and depression (38). Other unmeasured aspects of socioeconomic position (e.g. wealth), migration-related health determinants and adverse childhood exposures may also contribute to observed ethnic differences (39). Furthermore, attrition may introduce bias. Compared to the CCA sample, participants lost to follow-up had slightly greater age, BMI, as well as slightly fewer years of education, but were otherwise nearly identical in observable baseline characteristics (ESM 1).

Sleep disorders might be undertreated and underdiagnosed in the UK (40). Given the comparable effect size of poor sleep quality with traditional type 2 diabetes risk factors (6), identifying cost-effective and scalable early interventions to optimise sleep quality and duration show potential for type 2 diabetes prevention and for promoting population health.

In conclusion, our findings suggested that sleep quality, particularly snoring (a proxy for OSA) in middle-age is associated with the development of type 2 diabetes in later life, even after adjusting for traditional type 2 diabetes risk factors. This association was only found in South Asians but not among the Europeans and African Caribbeans. Findings do not suggest significant associations for difficulty falling asleep, early morning waking and waking up tired. This warrants further investigation into the apparent ethnic inequality and any sources of resilience due to changes in sleep pattern post-retirement. This study may inform strategies for type 2 diabetes prevention among South Asians through screening for snoring or OSA as risk factors for type 2 diabetes.

## Data Availability

The dataset analysed are available from the SABRE Study Group but restrictions apply to the availability of these data, which were used with permission for the current study, and so are not publicly available. Data are however available from the corresponding author upon reasonable request and with permission from the SABRE Study Group.

## Acknowledgements

The authors are grateful to the participants for their continued support and to the SABRE Study Group who contributed to the study design, study management and primary data collection.

## Contribution statement

CD and VG conceived the study idea and design. ZO, VG and TT contributed to the design of analyses. VG and ZO jointly performed the statistical analyses. ZO wrote the manuscript under the supervision of CD and VG. NC and TT contributed to the design and acquisition of the data. All authors contributed to the interpretation of the results, critically appraised the manuscript and approved the final draft. VG guarantees the work carried out, had access to all of the study data and takes responsibility for the integrity of the data and accuracy of the data analysis.

